# Development of a mathematical model to predict the health impact and duration of SARS-CoV-2 outbreaks on board cargo vessels

**DOI:** 10.1101/2021.11.03.21265201

**Authors:** Kok Yew Ng, Tudor A. Codreanu, Meei Mei Gui, Pardis Biglarbeigi, Dewar Finlay, James McLaughlin

**Affiliations:** Engineering Research Institute, Ulster University, Shore Road, Newtownabbey, BT37 0QB, UK; State Health Incident Coordination Centre, Department of Health Western Australia, Perth, Western Australia, Australia; Disaster Preparedness and Management Unit, Divisional Commander, Complex Medical Deployments, Department of Health Western Australia, Perth, Western Australia, Australia; School of Chemistry and Chemical Engineering, Queen’s University Belfast, David Keir Building, Stranmillis Road, Belfast, BT9 5AG, UK; Flowminder Foundation, Regus, Cumberland House, Grosvenor Square, Southampton, SO15 2BG, UK; Electrical and Computer Systems Engineering, School of Engineering, Monash University Malaysia, Jalan Lagoon Selatan, Bandar Sunway, 47500, Selangor, Malaysia

**Author notes:** Contributing authors. These authors contributed equally to this work (Kok Yew Ng, Tudor A. Codreanu).

**Keywords:** COVID-19, Coronavirus, Mathematical modelling, Cargo vessels, Western Australia

## Abstract

The Coronavirus Disease (COVID-19) pandemic has brought significant impact onto the maritime activities worldwide, including disruption to global trade and supply chains. The ability to predict the evolution and duration of a COVID-19 outbreak on cargo vessels would inform a more nuanced response to the event and provide a more precise return-to-trade date. A SEIQ(H)R (Susceptibility-–Exposed–Infected—Quarantine-–(Hospitalisation)—Removed/Recovered) model is developed and fit-tested to simulate the transmission dynamics of COVID-19 on board cargo vessels of up to 60 crew. Due to specific living and working circumstances on board cargo vessels, instead of utilising the reproduction number, we consider the highest fraction of crew members who share the same nationality to quantify the transmissibility of the disease. The performance of the model is verified using case studies based on data collected during COVID-19 outbreaks on three cargo vessels in Western Australia during 2020. The simulations show that the model can forecast the time taken for the transmission dynamics on each vessel to reach their equilibriums, providing informed predictions on the evolution of the outbreak, including hospitalisation rates and duration. The model demonstrates that (a) all crew members are susceptible to infection; (b) their roles on board is a determining factor in the evolution of the outbreak; (c) an unmitigated outbreak could affect the entire crew and continue on for many weeks. The ability to model the evolution of an outbreak, both in duration and severity, is essential to predict outcomes and to plan for the best response strategy. At the same time, it offers a higher degree of certainty regarding the return to trade, which is of significant importance to multiple stakeholders.

## 1 Introduction

Since the declaration of the Coronavirus Disease 2019 (COVID-19) pandemic by the World Health Organisation (WHO) (World Health Organization, 2020b), merchant vessels worldwide have been affected by Severe Acute Respiratory Syndrome Coronavirus 2 (SARS-CoV-2) outbreaks and ports remain a high-risk entry point for the infection (Codreanu et al, 2021b; Codreanu and Armstrong, 2021; Codreanu et al, 2021a). The spread of infectious diseases amongst crew on board cargo vessels is facilitated by several factors including a high population density in frequent direct contact areas, sharing of general amenities on board (Codreanu et al, 2021b; World Health Organization, 2020a), and interactions with shore-based maritime workers. Risk mitigation through international sanitation legislation requirements (World Health Organization, 2011a,b, 2016; EU SHIPSAN Act Joint Action, 2016; World Health Organization, 2005) and the measures mandated under the International Health Regulations (Codreanu et al, 2021b; Rocklöv et al, 2020; Tabata et al, 2020; Nakazawa et al, 2020) have decreased the transmissions of SARS-CoV-2 (Codreanu et al, 2021b,a; Rocklöv et al, 2020), but were unable to fully control outbreaks on board (Codreanu and Armstrong, 2021; Codreanu et al, 2021a; Walker et al, 2021).

Unlike cruise ships that accommodate very large numbers of passengers and crew (Codreanu et al, 2021a), cargo vessels operate with a comparatively much fewer crew (Codreanu et al, 2021b), and with very different work and social interaction patterns. Granting of pratique is based on providing objective and subjective data of the situation on board prior to arrival, of which assessment could be misleading (Codreanu and Armstrong, 2021). The declaration of an outbreak on board a cargo vessel has implications not only for the crew, but also for the vessel owner, shipping agent, operator, flag state, trade partners, and incident response agencies. Notwithstanding the threat to the crew’s health (Codreanu et al, 2021a; Schlaich et al, 2009), outbreak management not only has significant logistic and economic impact on the response agencies, but also on maritime transport and trade (Codreanu et al, 2021b; Jerome et al, 2017). In addition, the containment measures (quarantine, isolation, and sanitation), travel restrictions, and border closures (Australian Government, 2015) continue to make it increasingly difficult for ship operators worldwide to be granted pratique (Dujarric, 2015; Australian Government Department of Health, 2020), conduct trade, and change crew (Australian Government Department of Agriculture Water and Environment, 2020).

The Susceptibility–Infected–Removed (SIR) or Susceptibility–Exposed– Infected–Removed (SEIR) mathematical models have been used to describe the transmission dynamics of infectious diseases outbreaks (Kermack and McK-endrick, 1927) in which the epidemic is represented in a series of separate compartments or subpopulations. The SEIR model was used to predict the effectiveness of public health measures to control the COVID-19 outbreak on a passenger cruise ship (Rocklöv et al, 2020), and a modified SEIR model that includes hospitalisation, quarantine, and isolation was used to model other infectious disease epidemics (Safi and Gumel, 2010, 2011).

The generalised SEIQ(H)R model was developed and modified based on previous reported works (Safi and Gumel, 2010, 2011; Ng and Gui, 2020; Do et al, 2021) and represents the epidemic in separate compartments in cascade: susceptibility (S), exposed (E), infected (I), quarantine (Q), hospitalisation (H), and removed/recovered (R). This research is crucial towards the fight against the COVID-19 pandemic as we address the importance of modelling the transmission dynamics of SARS-CoV-2 amongst crew members on board cargo vessels, which has rarely been considered in published works thus far as most of them reported on passenger cruise ships (Rocklöv et al, 2020; Mizumoto and Chowell, 2020; Mizumoto et al, 2020).

The continuous advancement of knowledge on the clinical outcomes of COVID-19 and the role of associated factors such as age, co-morbidities, and ethnicity (Clift et al, 2020; Lighter et al, 2020) should underpin a more nuanced approach to managing outbreaks on cargo vessels in order to minimise the impact on crew health, maritime traffic, and trade. As a result, mathematical models can be used to provide an accurate and dynamic prediction of a COVID-19 outbreak progression in this setting.

Knowledge of the modelled evolution of the COVID-19 outbreak, both in duration and severity, is essential to predict outcomes and to plan for the most efficient and effective response strategies. Based on the Western Australian experience in managing COVID-19 outbreaks on board cargo vessels during 2020, here in this paper we describe the development of a mathematical model that could be used to inform the risk assessment of the population-specific epidemiological parameters for SARS-CoV-2 spread on similar vessels of up to 60 crew. This research is also important as the predictions from the model would help stakeholders in the analysis and understanding of the transmission dynamics of an infection disease on board a vessel during the earlier stages of a pandemic, especially when a vaccine has yet to be developed or that the availability of vaccine supplies for the general public is scarce. Hence, maritime operations during this period is most vulnerable to the impact of the pandemic with lack of quality testing facilities, as well as the impose of quarantines and closure of international borders and ports. Using this model, the responders to the outbreak will have a common understanding of the event evolution, including hospitalisation rates, as a function of the measures implemented.

This paper is organised as follows: Section 2 introduces the SEIQ(H)R mathematical model; Section 3 describes the data sources of the three cargo vessels used to verify and validate the proposed mathematical model; Section 4 verifies the proposed model and also provides some extensive results and discussions about the simulations; Section 5 presents the limitations of this research; and Section 6 concludes the paper.

## 2 Methodology

The SEIQ(H)R model was developed utilising data recorded during quarantine measures and hospitalisations on board the vessels, and the following assumptions: (i) absence of vaccinations against SARS-CoV-2, (ii) absence of confirmed history of previous COVID-19 infections, and (iii) absence of non-COVID-19-related deaths occurring on the vessels. Hence, the model can be expressed using

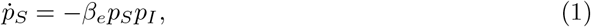

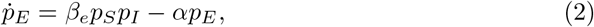

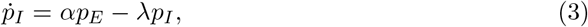

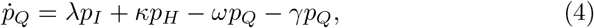

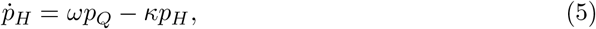

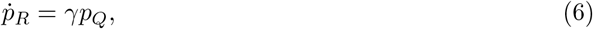

where *p*_*S*_, *p*_*E*_, *p*_*I*_, *p*_*Q*_, *p*_*H*_, and *p*_*R*_ represent the susceptible (S), exposed (E), infectious (I), quarantined (Q), hospitalised (H), and removed/recovered (R) subpopulations, respectively. The parameter *β*_*e*_ denotes the rate of transmission per S-I contact, *α* the rate of an exposed person becoming infectious, *λ* the rate of which an infectious person is quarantined, *ω* the rate of which a quarantined person is hospitalised, *γ* the rate of which a quarantined person recovers, and *κ* is the rate of which a hospitalised person is discharged and completes their quarantine.

Thus, the incubation time can be written as *τ*_*inc*_ = 1*/α*, the time from onset to being quarantined is *τ*_*infQ*_ = 1*/λ*, the time spent in quarantine before recovery is *τ*_*quarR*_ = 1*/γ*, the time spent in quarantine before being hospitalised is *τ*_*quarH*_ = 1*/ω*, and the hospitalisation time is *τ*_*hosp*_ = 1*/κ*. The total number of crew members on the vessel, *p*_*N*_, can be computed such that *p*_*N*_ = *p*_*S*_ + *p*_*E*_ + *p*_*I*_ + *p*_*Q*_ + *p*_*H*_ + *p*_*R*_.

Given the crew size, the transmission dynamics and spread of the infectious disease cannot be accurately quantified using the reproduction number that is commonly applied to studies involving large populations, i.e. a country or a geographical region. Thus, instead of using the parameter *β*_*e*_ (or its equivalent mathematical representation in other existing models) to compute the reproduction number (Safi and Gumel, 2010, 2011; Ng and Gui, 2020; Do et al, 2021), its purpose here is to inform the transmission dynamics of SARS-CoV-2 by considering the majority of crew members who share the same nationality. This information is estimated critical as crew who originate from the same country often share similar language and culture, thus have increased social interactions on board, which in turn would reasonably be expected to increase the rate of transmission. Therefore, in this paper, *β*_*e*_ is expressed using

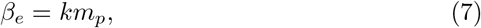

where *m*_*p*_ represents the highest fraction of crew who originate from the same country and *k* is a scalar coefficient. Figure 1 shows the block diagram of the SEIQ(H)R model in equations (1)–(6).

**Fig. 1.**
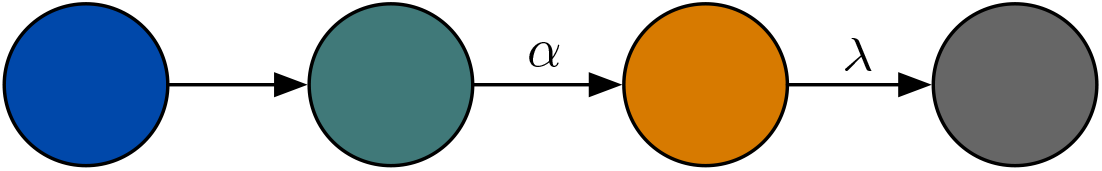
The block diagram of the SEIQ(H)R model in equations (1)–(6).

## 3 Data Sources

The data used are based on the records of COVID-19 outbreaks declared on the cargo vessels MV Al Kuwait, MV Al Messilah, and MV Patricia Oldendorff during 2020 in Western Australia.

The data collection, analysis, storage, and reporting were conducted in line with the WHO Ethical Standards for Research During Public Health Emergencies (COVID-19), the WHO Guidance for Managing Ethical Issues in Infectious Diseases Outbreaks, and the WHO Guidelines on Ethical Issues in Public Health Surveillance. Ethics approval was not required for this investigation as it was conducted as part of the public health response to the outbreaks of COVID-19, which is a notifiable infectious disease under the Western Australia Public Health Act 2016. The release of data not already in the public domain has been granted by the Western Australia Department of Health Public Health Emergency Operations Centre Data Custodian. The Western Australian Department of Health Human Research Ethics Committee (HREC) has determined that the routine public health investigative work relating to a series of outbreaks of infectious disease that involved aggregate data, in accordance with Section 5.1.8 of the National Statement, involves only negligible risk (i.e. where there is no foreseeable risk or harm or discomfort, and any foreseeable risk is no more than inconvenience) and therefore does not need to be reviewed by the HREC.

## 4 Results and Discussion

In aggregate, 63 out of 122 crew were diagnosed with COVID-19 (51.64%), and three (4.76%) of the cases required hospitalisation, but not mechanical ventilatory support. Table 1 summarises the data from the three vessels.

**Table 1.** Summarised data of the three vessels.

| Description | Value |
| --- | --- |
| **<u>Vessel 1: MV Al Kuwait</u>** |  |
| Total crew members | 48 |
| <u>Crew members by nationalities</u> |  |
| Philippines | 32 (66.67%) |
| Croatia | 10 (20.83%) |
| India | 3 (6.25%) |
| Australia | 2 (4.17%) |
| Tanzania | 1 (2.08%) |
| Total confirmed cases | 21 |
| Total hospitalised | 2 |
| Mean quarantine days | 16.4 |
| Mean hospitalisation days | 2 |
| **<u>Vessel 2: MV Al Messilah</u>** |  |
| Total crew members | 53 |
| <u>Crew members by nationalities</u> |  |
| Bangladesh | 37 (69.81%) |
| India | 7 (13.21%) |
| Sri Lanka | 4 (7.54%) |
| Pakistan | 3 (5.66%) |
| Australia | 1 (1.89%) |
| Syria | 1 (1.89%) |
| Total confirmed cases | 25 |
| Total hospitalised | 1 |
| Mean quarantine days | 10.5 |
| Mean hospitalisation days | 2 |
| **<u>Vessel 3: MV Patricia Oldendorff</u>** |  |
| Total crew members | 21 |
| <u>Crew members by nationalities</u> |  |
| Philippines | 20 (95.24%) |
| Poland | 1 (4.76%) |
| Total confirmed cases | 17 |
| Total hospitalised | 0 |
| Mean quarantine days | 9.8 |
| Mean hospitalisation days | — |

Simulations to verify the model were carried out using MATLAB/Simulink R2021a. Given the data in Table 1, and to ensure that the model can be generalised, the parameters of the model in (1)–(6) for the three cargo vessels are set as shown in Table 2. The parameter *m*_*p*_ is determined by the highest fraction of nationality in each vessel. The scalar coefficient *k* is a design freedom and is set to 1.15 for all three vessels to indicate that it does not need to be tuned and can be applied generally to cargo vessels of similar settings. The rate of an exposed crew becoming infectious *α* is the reciprocal of the incubation period and is set to 1*/*5 in accordance to Flaxman et al (2020). The remaining parameters in Table 2, i.e. *λ, γ, ω*, and *κ*, are set using the mean values obtained from the data.

**Table 2.** Parameters settings of the SEIQ(H)R model for the three cargo vessels used in the case studies.

| Parameter | Description | MV Al Kuwait | MV Al Messilah | MV Patricia Oldendorff |
| --- | --- | --- | --- | --- |
| $m_p$ | Highest fraction of crew who share the same nationality | 0.67 | 0.70 | 0.95 |
| $k$ | Scalar coefficient | 1.15 | 1.15 | 1.15 |
| $\alpha$ | Rate of an exposed crew becoming infectious (5 days) | 1/5 | 1/5 | 1/5 |
| $\lambda$ | Rate of an infectious crew being quarantined (5 days) | 1/5 | 1/5 | 1/5 |
| $\gamma$ | Rate of a quarantined crew recovers (95%, 15 days) | $0.95 \times 1/15$ | $0.95 \times 1/15$ | $0.95 \times 1/15$ |
| $\omega$ | Rate of a quarantined crew is hospitalised (5%, 2 days) | $0.05 \times 1/2$ | $0.05 \times 1/2$ | $0.05 \times 1/2$ |
| $\kappa$ | Rate of a hospitalised crew is discharged (2 days) | 1/2 | 1/2 | 1/2 |

The convergence between the results of the proposed model and the observed data for each vessel are presented in Figure 2, which shows that the simulations can forecast the time for the transmission dynamics on each vessel to reach their equilibriums. Thus, these results are able to provide informed predictions on the evolution of the outbreak on board that would be useful to determine if and when the vessel can be granted pratique to conduct trade and crew exchange.

**Fig. 2.**
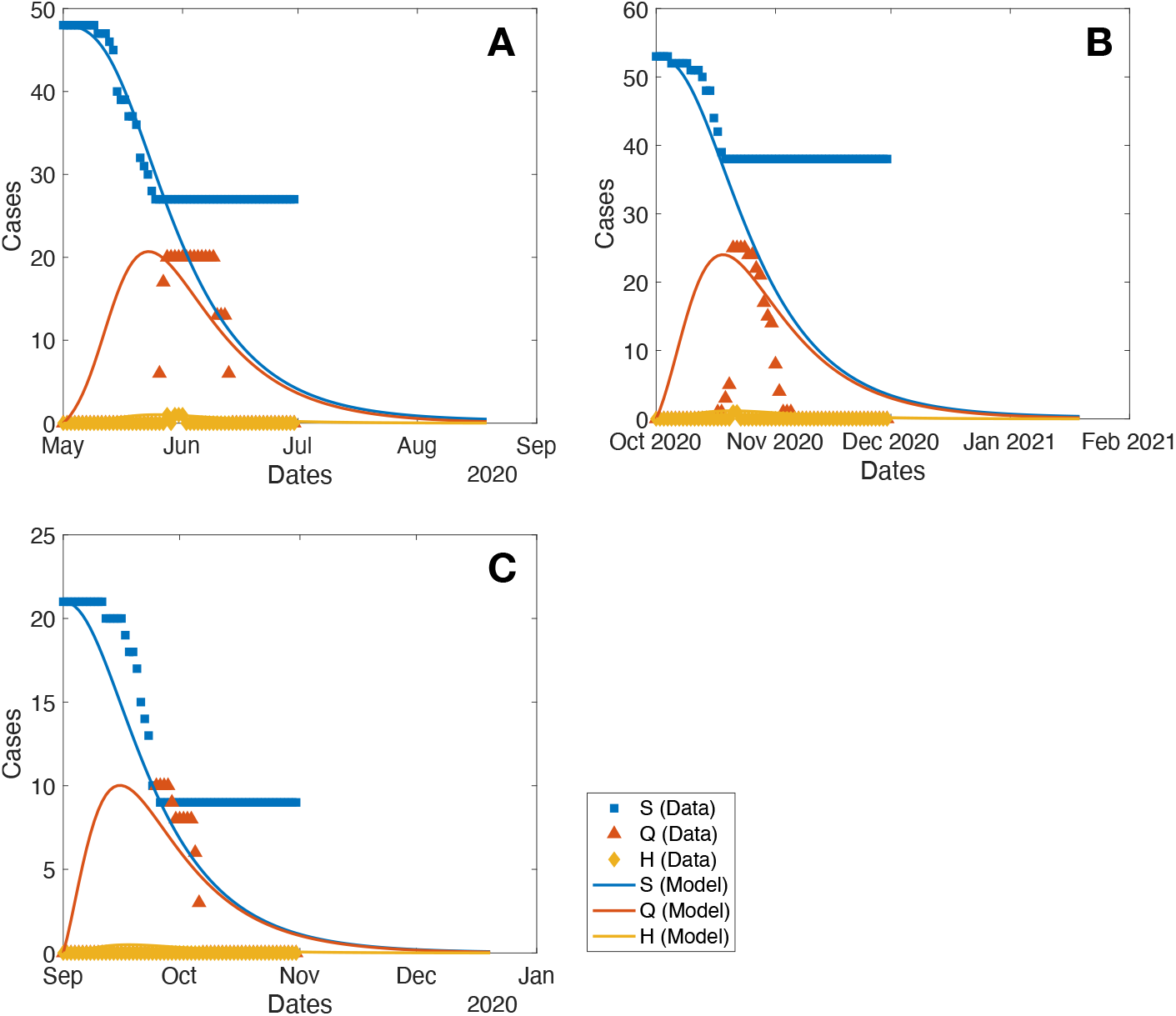
Data and model plots for the S, Q, and H compartments of vessels A) MV Al Kuwait, B) MV Al Messilah, and C) MV Patricia Oldendorff.

The model also suggests that all crew members should be considered susceptible to infection and that, if left unmitigated, the transmission of the infection on board can

(a) affect the entire crew, potentially crippling the safety and security of the vessel by breaching the mandated Minimum Safe Manning (Codreanu et al, 2021b), and
(b) continue on for many weeks, contrary to estimations declared by some operators (Codreanu and Armstrong, 2021)

The very low number and short duration of hospital admissions could reflect the relative absence of pre-existing or underlying health risk factors in the individuals affected, which supports a nuanced approach to the management of the outbreak. However, the risks of severe disease and death are not null, and a clinical deterioration at sea can have catastrophic consequences to the individuals, the larger crew, and the vessel. This is further complicated by the default absence of medical facilities, monitoring equipment, and qualified medical personnel on board cargo vessels.

It was also found that crew members’ roles on board is a determining factor in the evolution of the outbreak (Figure 3). This could be due to the characteristics of the accommodation structure on cargo vessels that include common predicted intersection points (e.g. mess rooms and catering staff, as well as shared facilities such as showers and toilets) irrespective of the technical segregation between officers and ratings, in both the deck and engine groups. This is evidenced in Figure 3A where only three groups of crew occupations have tested positive. Given that the Deck Rating crew have limited physical interactions with the Engine Rating crew and also that they work in different sections of the vessel, it can be assumed that the Catering Ratings are a common intersection point in the transmission of the infection. Figure 3 shows that a higher *m*_*p*_ value would lead to a higher total number of positive cases.

The Maritime Labour Convention, 2006 (International Labour Organization, 2006) requires that separate mess room facilities are provided to crew members of different ranking; (i) master and officers, and (ii) petty officers and other crew. Thus, officers and higher ranked crew members would use Mess Room 1, and the rest of the crew, Mess Room 2. However, all hot meals are prepared and delivered by the same catering staff. Figure 4 demonstrates that, on each vessel, there were higher number of positive cases recorded in crew from the same nationality (or closely related cultures) with the catering staff. These observations agree with the hypothesis that the more similar the culture and language, irrespective of nationality, the more social interactions occur on board, which, in turn, increases the transmission of the virus.

**Fig. 3.**
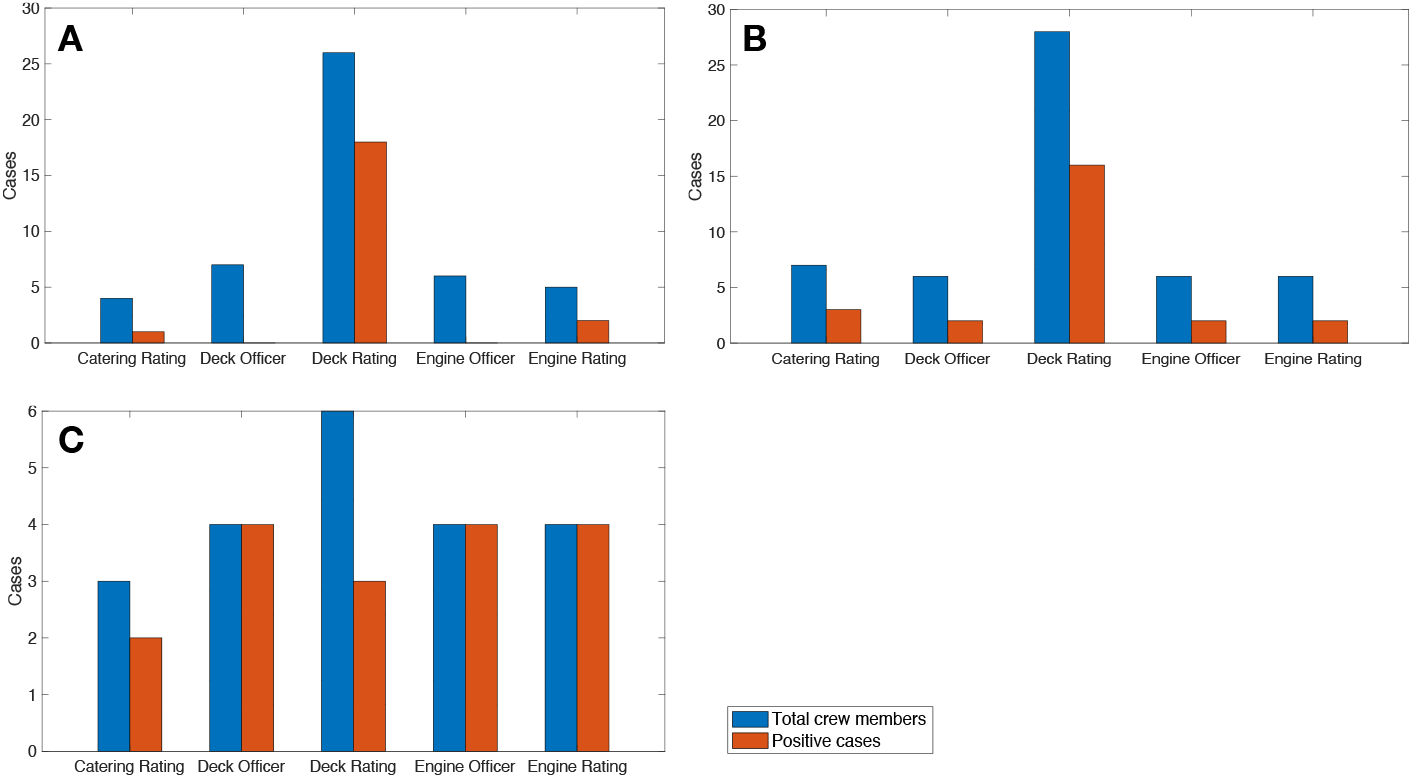
Cases by crew occupation (officers and ratings) for vessels A) MV Al Kuwait, B) MV Al Messilah, and C) MV Patricia Oldendorff. The Catering Ratings are a common intersection point for all vessels.

**Fig. 4.**
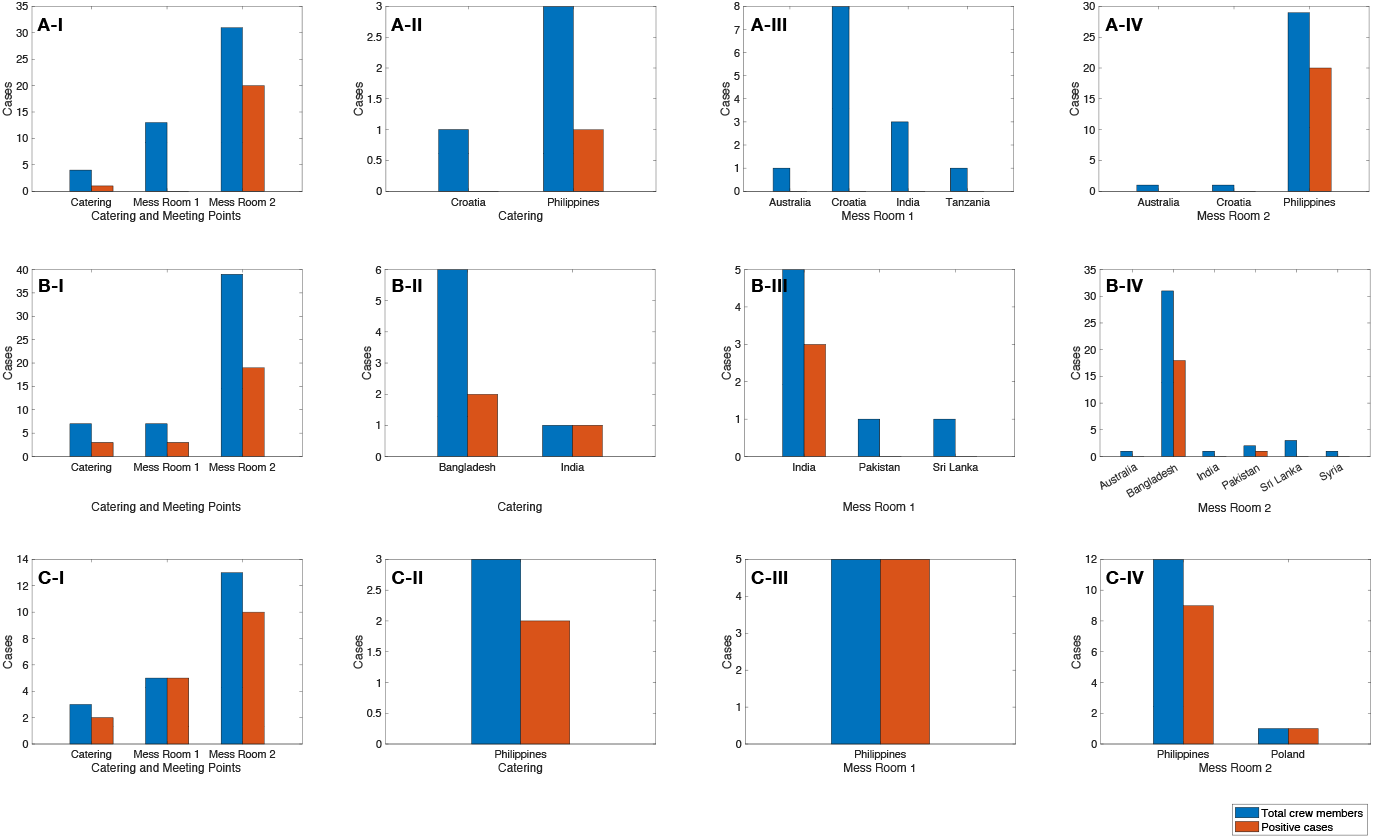
Total crew members (blue) and positive cases (red) in Catering, Mess Room 1, and Mess Room 2 intersection points for vessels A) MV Al Kuwait, B) MV Al Messilah, and C) MV Patricia Oldendorff, respectively. Subplots (I) show the collective data for each vessel whilst subplots (II)–(IV) show the distribution of each intersection point by nationality.

## 5 Limitations

The proposed model fits the data collected during the management of the three COVID-19 outbreaks described and is limited to a typical cargo vessel crew size of no more than 60 crew members. More data would be required to ascertain that the observed model and data convergence can be generalised to larger crew sizes. The model has not been calibrated for complex individualist versus collectivist cultural aspects of the crew, nor for social interaction patterns on board as these are largely superseded by clear hierarchical command and control relationships as well as work-related intersection points. Therefore, further division of the crew in subclasses of susceptible individuals has not been deemed necessary as it is not expected to alter the modelling. In addition, population of passengers are not considered in the mathematical model as the operations of cargo vessels differ significantly from passenger cruise vessels.

Also, vaccines against SARS-CoV-2 were not available at the time of the study. Whilst noting that the main aim of vaccines is to reduce the risk of severe disease and death, and not necessarily the susceptibility to infection, vaccination status is crucial in the modelling design as the predictions would help stakeholders in the analysis and understanding of the transmission dynamics of an infection disease on board a cargo vessel. Early stages of any outbreaks with little known pathogens and adequate mitigation strategies are also the periods of which maritime operations are most affected: absence of readily accessible testing platforms and/or facilities, impost of quarantine of varying lengths, and unpredictable duration of closure of international borders and ports. Modelling works may not be as relevant during the later stages of the pandemic when most population would have been protected by the development of vaccines or natural herd immunity. However, the continuous emergence of variants of concern which may result in reinfections or evade the enhanced immunity provided by current vaccines, and the inequal availability of vaccines around the world provides enough reasons for the enduring validity of this mathematical concept.

The model does not consider individual’s past COVID-19 illness as quality information is usually not available at the time of the outbreak declaration. Since gathering and verifying the crew’s previous COVID-19 infection data may prove logistically challenging where reliable information is lacking, serological determination may be required that prolongs the timeline to verification. Further refining of the model could be undertaken by incorporating emerging evidences regarding the protection to SARS-CoV-2 (re)infection in vaccinated individuals.

## 6 Conclusion

COVID-19 outbreaks on cargo vessels continue to present significant economic, trade, and health impacts to countries worldwide (United Nations Conference on Trade and Development (UNCTAD), 2021; Goodman, 2022; Murray et al, 2022). The ability to model the evolution of an outbreak on board typical crew-sized cargo vessels, both in duration and severity, is essential to predict outcomes and to plan for the best response strategies. The proposed SEIQ(H)R model has provided a good fit for the outbreaks on three cargo vessels in Western Australia in 2020. Further research is required to determine its generalisability onto other classes and categories of cargo vessels, whilst also focussing on new and emerging SARS-CoV-2 variants, as well as previous COVID-19 disease and/or vaccination status.

## Data Availability

All data produced in the present study are available upon reasonable request to the authors

## Acknowledgments

To the crew and Masters of the vessels MV Al Kuwait, MV Al Messilah, MV Patricia Oldendorff, and MV Key Integrity — our heartfelt and sincere recognition for your compliance during the challenges of quarantine.

The authors would also like to acknowledge the contributions of the representatives of the following State agencies who have made possible the execution of the Western Australian State COVID-19 outbreak quarantine sequence on board these vessels: Western Australian Department of Health, Western Australian Medical Assistance Team, Australian Border Force, Western Australian Police Force, Department of Fire and Emergency Services, Fremantle Port, Pilbara Port, Western Australian Country Health Services, PathWest, Royal Flying Doctor Service, Australian Defence Force, Australian Maritime Safety Authority, Department of Agriculture, Water and the Environment, Department of Foreign Affairs and Trade. Special acknowledgments are due to the in-country representatives of the affected vessels (shipping agents).

## Declarations

